# Health seeking behaviors of patients with acute respiratory infections during the outbreak of novel coronavirus disease 2019 in Wuhan, China

**DOI:** 10.1101/2020.05.05.20091553

**Authors:** Juan Yang, Hui Gong, Xinhua Chen, Zhiyuan Chen, Xiaowei Deng, Mengcen Qian, Zhiyuan Hou, Marco Ajelli, Cecile Viboud, Hongjie Yu

## Abstract

We conducted two surveys to evaluate the health-seeking behaviors of individuals with acute respiratory infections (ARI) during the COVID-19 outbreak in Wuhan, China. Among 351 participants reporting ARI (10.3%, 351/3,411), 36.5% sought medical assistance. Children were more likely to seek medical assistance than other age groups (66.1% vs. 28.0%-35.1%).

## Introduction

The novel coronavirus disease 2019 (COVID-19) emerged in Wuhan, Hubei province, China in December 2019 and rapidly spread across the world[1, 2]. During emergency situations, rapid measurements of changes in health-seeking behavior is critical to understand healthcare utilization. Moreover, estimating the disease burden and clinical severity of COVID-19 is key to identify appropriate intervention strategies and allocate healthcare resources. However, quantifying these metrics while a pandemic is unfolding is challenging due to limitations in passive surveillance. Here, we assess the health seeking behavior of residents with acute respiratory infections (ARI) during the outbreak of COVID-19 between December 2019 and March 2020 in Wuhan, China.

## Methods

Between March 10 and 24, 2020, two population-based surveys were conducted in Wuhan to understand the health seeking behaviors of patients suffering from ARI during the COVID-19 outbreak. The surveys included: i) a telephone-and-online survey of 2,595 adults (individuals aged ≥18 years); ii) an online survey of 816 children (individuals aged <18 years). The study participants were current residents who had lived in Wuhan for at least three months before the date of the survey. Based on the assumption that 6% of the population would have ARI [3], we calculated that a minimum sample size of 784 participants per age group (i.e., 3-17, 18-39, 40-59, 60 years of age) would allow a healthcare seeking proportion of 60% to be estimated, with a statistical significance level of 5%, and 14% of marginal error.

The telephone-and-online survey among adults was conducted in two steps. First, we randomly dialed Wuhan mobile phone numbers to invite the participants through a computer-assisted interviewing system. An overview of the study was provided at the beginning of each call. We attempted to contact each generated number twice a day at different hours and one more time on the following day; if no contact was established after the third call, the number was classified as invalid or unreachable. Second, we sent a telephone message with the online link of our questionnaire to the identified participants, who were asked to complete the questionnaire on their own. Of 48,965 calls answered, 2,858 persons (5.8%) completed the questionnaires (Figure S1 in Appendix).

The online survey of children was commissioned to the ePanel data company, which has a national online survey library with emails and household information registered. An online link to our questionnaire was provided to Wuhan families with children. The questionnaire was answered by the parents of recruited children. Among 35,770 email invitations, 933 persons (2.6%) completed the questionnaires (Figure S1 in Appendix).

Random dials/emails with proportional quota sampling were used to ensure that respondents were demographically representative of the general population, with quotas based on sex (female-to-male 1:1). Characteristics of respondents were collected, including age and sex. We also obtained the history of ARI for respondents in the last three months (presence of fever and/or any respiratory symptoms, e.g., cough and sore throat), and if they sought medical assistance for these symptoms. Questionnaires completed in less than 2 minutes and containing logical errors were excluded from the analysis (Figure S1 in Appendix). The full questionnaires are reported in Appendix.

To estimate the proportion of individuals with ARI and their probability of seeking medical assistance, participants’ ages were weighted to match the age structure of the Wuhan population. This corrected the underrepresentation of individuals aged 60+ years. Chi-square test and Fisher exact test were used for binary variables. Two-sided P values <0.05 were considered to indicate statistical significance. Binomial distributions were used to estimate the 95%CIs for binary variables.

The surveys were approved by the Institutional review board from School of Public Health, Fudan University (IRB#2020-01-0801). Informed consent was obtained from all respondents/parents of children before surveys.

## Results

A total of 3,411 study participants were included in our analysis. 49.6% (1,691/3,411) were female. Of them, 351 participants (10.3%, 95%CI 9.3%-11.4%) reported ARI during the COVID-19 outbreak in Wuhan, with higher proportions of ARI observed in adults than children (9.3%-12.5% depending on the age group vs. 6.9%, χ^2^=17.57, p<0.001). Among ARI cases, 36.5% (95% CI 31.5%-41.8%) sought medical care. No significant difference of health seeking behaviors was observed between male and female. (Figure S2 in Appendix) Children were more likely to seek medical assistance than other age groups (66.1% vs. 28.0%-35.1%, χ^2^=26.50, p<0.001). (see Table). Adjusting for the age structure of the Wuhan population, the overall proportion of patients with ARI was 10.5% (95%CI 9.3%-11.9%), and the adjusted proportion of seeking medical assistance for ARI was 35.4% (95%CI 28.4%-43.9%).

A small proportion of patients (16.4%, 95%CI 10.7%-24.2%) with ARI sought medical care in private clinics, while around 30% visited community-based health service centers or different types of hospitals. Of the patients who sought medical care, children aged 3-17 years and adults aged 60+ years were more likely to be hospitalized and admitted to ICU. Among children who did not seek medical care, 54.5% (95%CI 22.9%-82.9%>) reported to be afraid of nosocomial infection. Among adults who did not seek medical care, 51.6%-54.2% reported mild illness as a reason for their behavior (see Table).

## Discussion

This population-based study provides valuable insights in the health seeking behaviors of ARI patients during the COVID-19 outbreak (from December 2019 through to March 2020) in Wuhan. Although adults were more likely to report ARI than children aged 3-17 years, a smaller proportion of adults sought medical assistance. Among the reasons for not seeking medical care, parents reported concerns with nosocomial infections for their sick children, while experiencing mild illness was linked to lack of seeking care in adults. Our findings highlight the possible relevance of self-isolation at home and/or at designated medical facilities to manage mild cases and lower the load on hospitals [4].

The proportion of seeking medical assistance of ARI patients during COVID-19 outbreak in Wuhan was lower than that during 2009 influenza pandemic (35.4% vs. 46.1%) [3]. The health seeking behavior of typical patients with ARI may differ from that of COVID-19 cases. We asked adult participants if they were diagnosed as laboratory-confirmed or probable COVID-19 cases. Only 2.2% of them (57/2,595) self-reported to be a laboratory-confirmed or probable COVID-19 case. Among COVID-19 patients, 35.1% (20/57) did not answer the questions regarding healthcare seeking behavior. Hence there was not enough data to compare healthcare seeking behavior between COVID-19 cases and those with typical respiratory diseases. Additionally, the number of questionnaires completed by participants aged 60+ years was relatively small (n=361), probably due to the hurdles encountered by this segment of the population to fill an online survey. Therefore, we adjusted the overall proportion of ARI patients, and the proportion of seeking medical assistance, by adjusting for the actual age structure of Wuhan population.

A range of studies have quantified the clinical severity of COVID-19, particularly as regards the case and infection fatality ratios[5-8]. However, these estimates were mainly based on laboratory-confirmed cases captured by surveillance system, which may overestimate COVID-19 severity. In fact, we found that the majority of mild cases did not seek medical assistance and thus were hardly detectable by surveillance systems. The number of COVID-19 cases in the community level remains unclear due to changes in health seeking behaviors during an outbreak; an assessment of the full severity pyramid of COVID-19 is lacking. While several studies have assessed the healthcare demand of the COVID-19 pandemic on hospital beds, ICU beds and ventilators [9, 10], no study has evaluated the demand on outpatient facilities. Despite the aforementioned limitations, our study provides key metrics that can be instrumental to fill this gap.

## Conclusions

This population-based study demonstrates that the majority of patients with ARI symptoms did not seek medical assistance during the COVID-19 outbreak in Wuhan. These findings may be used to refine the estimates of disease burden and clinical severity of COVID-19, and the health demand associated with this pandemic, and to plan for health resources allocation.

## Data Availability

The raw/processed data required to reproduce these findings cannot be shared at this time as the data also forms part of an ongoing study.

## Notes

### Contributions

H.Y. conceived, designed and supervised the study. J.Y., H.G., X.C., Z.C., X.D., M.Q., and Z.H. participated in data collection. J.Y., H.G., X.C., and Z.C. analyzed the data, and prepared the table and figure. J.Y. prepared the first draft of the manuscript. M.A., C.V. and H.Y. commented on the data and its interpretation, revised the content critically. All authors contributed to review and revision and approved the final manuscript as submitted and agree to be accountable for all aspects of the work.

### Funding

The study was supported by grants from the National Science Fund for Distinguished Young Scholars (No. 81525023), Key Emergency Project of Shanghai Science and Technology Committee (No. 20411950100), National Science and Technology Major Project of China (No. 2018ZX10201001-010, No. 2018ZX10713001-007, No. 2017ZX10103009-005).

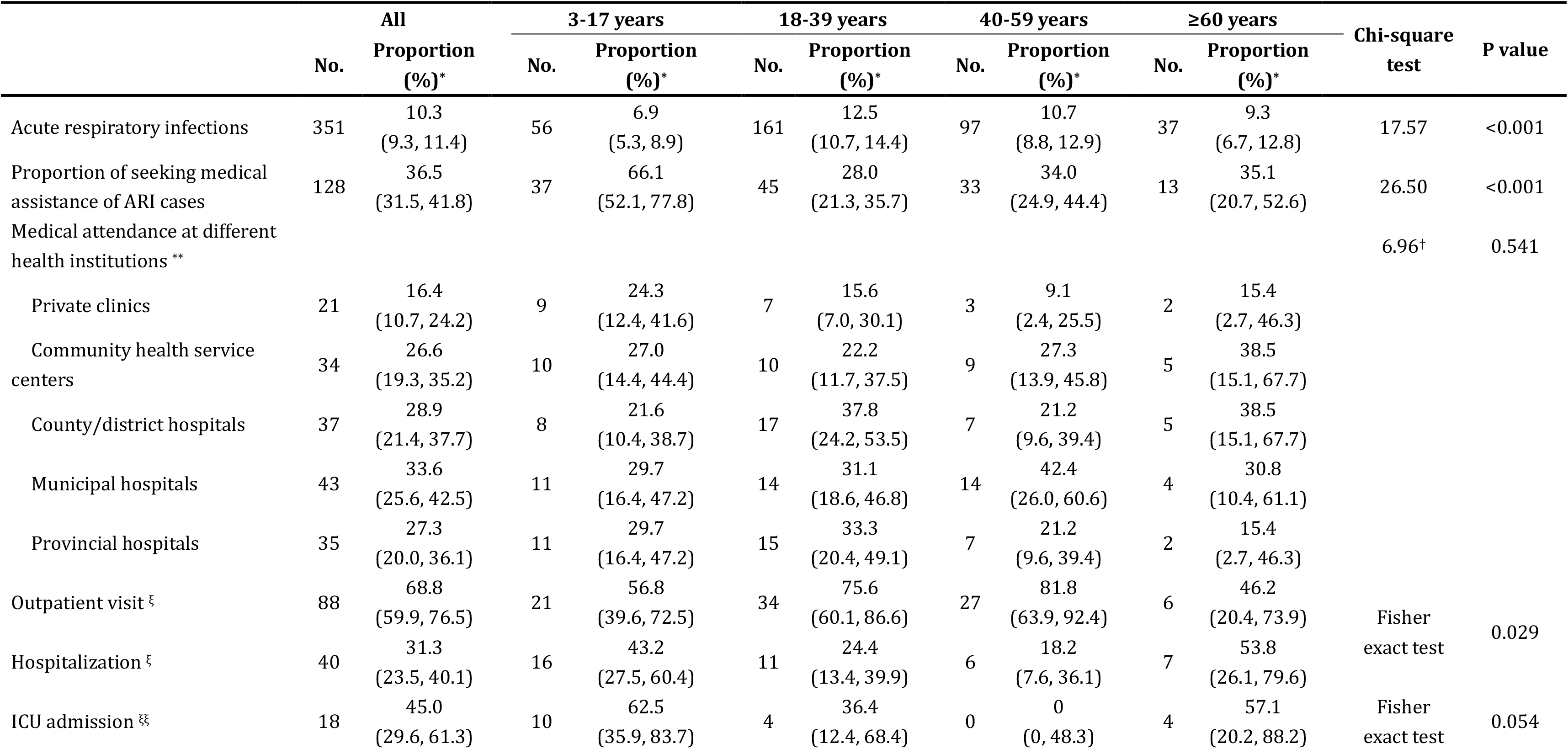
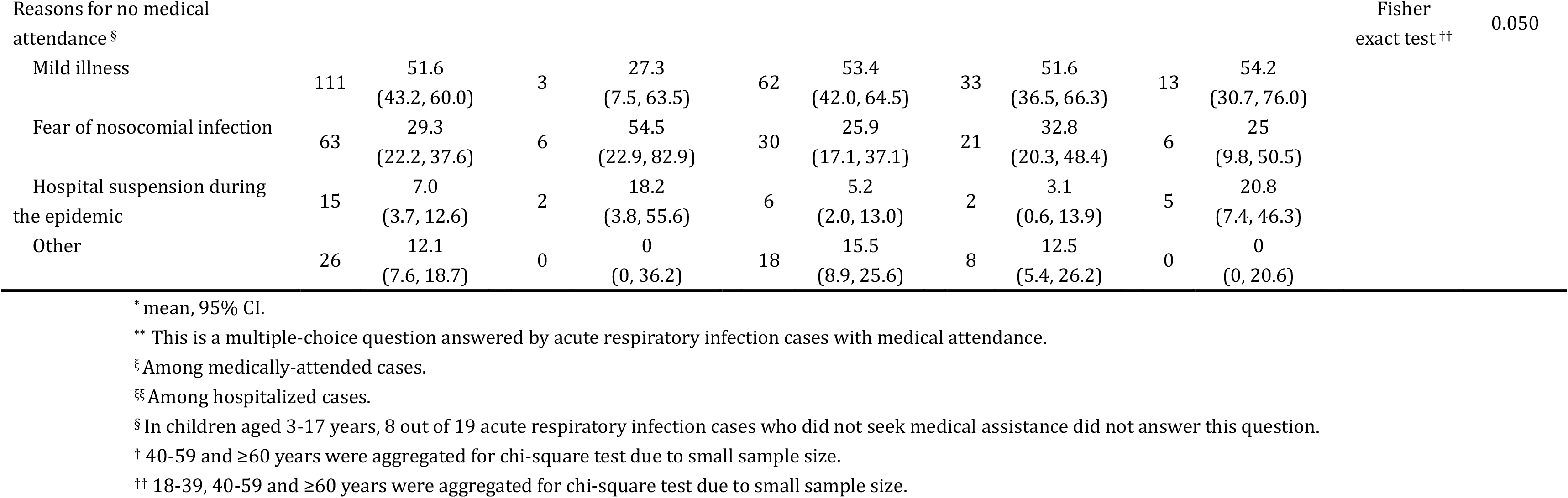
Table Health seeking behaviors of residents with acute respiratory infections during the COVID-19 pandemic in Wuhan, stratified by age groups

